# Clinical phenotype and management of sound-induced pain: Insights from adults with pain hyperacusis

**DOI:** 10.1101/2024.06.19.24309185

**Authors:** Kelly N. Jahn, Sean Takamoto Kashiwagura, Muhammad Saad Yousuf

**Affiliations:** Department of Speech, Language, and Hearing, The University of Texas at Dallas, Richardson, TX, USA; Callier Center for Communication Disorders, The University of Texas at Dallas, Dallas, TX, USA; Department of Neuroscience and Center for Advanced Pain Studies, The University of Texas at Dallas, Richardson, TX, USA

## Abstract

Pain hyperacusis, also known as noxacusis, causes physical pain in response to everyday sounds that do not bother most people. How sound causes excruciating pain that can last for weeks or months in otherwise healthy individuals is not well understood, resulting in a lack of effective treatments. To address this gap, we identified the most salient physical and psychosocial consequences of debilitating sound-induced pain and reviewed the interventions that sufferers have sought for pain relief to gain insights into the underlying mechanisms of the condition. Adults (*n* = 32) with pain hyperacusis attended a virtual focus group to describe their sound-induced pain. They completed three surveys to identify common symptoms and themes that defined their condition and to describe their use of pharmaceutical and non-pharmaceutical therapies for pain relief. All participants endorsed negative effects of pain hyperacusis on psychosocial and physical function. Most reported sound-induced burning (80.77%), stabbing (76.92%), throbbing (73.08%), and pinching (53.85%) that occurs either in the ear or elsewhere in the body (i.e., referred pain). Participants reported using numerous pharmaceutical and non-pharmaceutical interventions to alleviate their pain with varying degrees of pain relief. Benzodiazepines and nerve blockers emerged as the most effective analgesic options while non-pharmaceutical therapies were largely ineffective. Symptoms of pain hyperacusis and therapeutic approaches are largely consistent with peripheral mechanistic theories of pain hyperacusis (e.g., trigeminal nerve involvement). An interdisciplinary approach to clinical studies and the development of animal models is needed to identify, validate, and treat the pathological mechanisms of pain hyperacusis.

## INTRODUCTION

Approximately 15% of adults [6] and 17% of children [15] suffer from hyperacusis, a disorder characterized by an inability to tolerate ordinary sounds that do not bother most people [19]. Some individuals perceive everyday sounds to be excessively loud (i.e., *loudness hyperacusis*), whereas others experience physical pain in response to sounds (i.e., *pain hyperacusis* or *noxacusis*) [19]. The relative prevalence of these two hyperacusis subtypes is unknown, but a recent survey demonstrated that 62.5% of adults with hyperacusis experience some level of sound-induced physical pain [21]. Individuals with pain hyperacusis often present with a relatively severe clinical phenotype, wherein they report frequent “setbacks” (i.e., symptom exacerbations in response to a trigger sound) and reduced benefit from behavioral interventions (e.g., sound therapy) [21]. Despite the severity of pain hyperacusis, healthcare professionals have limited tools to diagnose, monitor, and treat the condition [2] and they do not widely recognize the pain hyperacusis phenotype [7].

A primary reason for the lack effective clinical tools for pain hyperacusis is that the underlying mechanisms of sound-induced pain remain elusive [6]. Existing theories of the neural underpinnings of pain hyperacusis implicate structures ranging from the middle ear to the inner ear to the central auditory pathway (**Figure 1**). Middle ear models of sound-induced pain broadly suggest that overload, damage, or myoclonus of the tensor tympani muscle can irritate the trigeminal nerve and cause pain in or near the ear [12,20]. Others have shown that Type-II cochlear afferents share similar morphological and neurochemical properties with nociceptive somatic C fibers and may transmit damage-evoked pain signals from the inner ear to the central nervous system (CNS) [4,10,22]. Finally, a large body of basic science literature demonstrates that damage to the peripheral auditory system leads to a cascade of downstream changes that elicit abnormally elevated neural activity along the central auditory pathway and which may lead to perceptual hypersensitivity in rodents [1].

**Figure 1.**
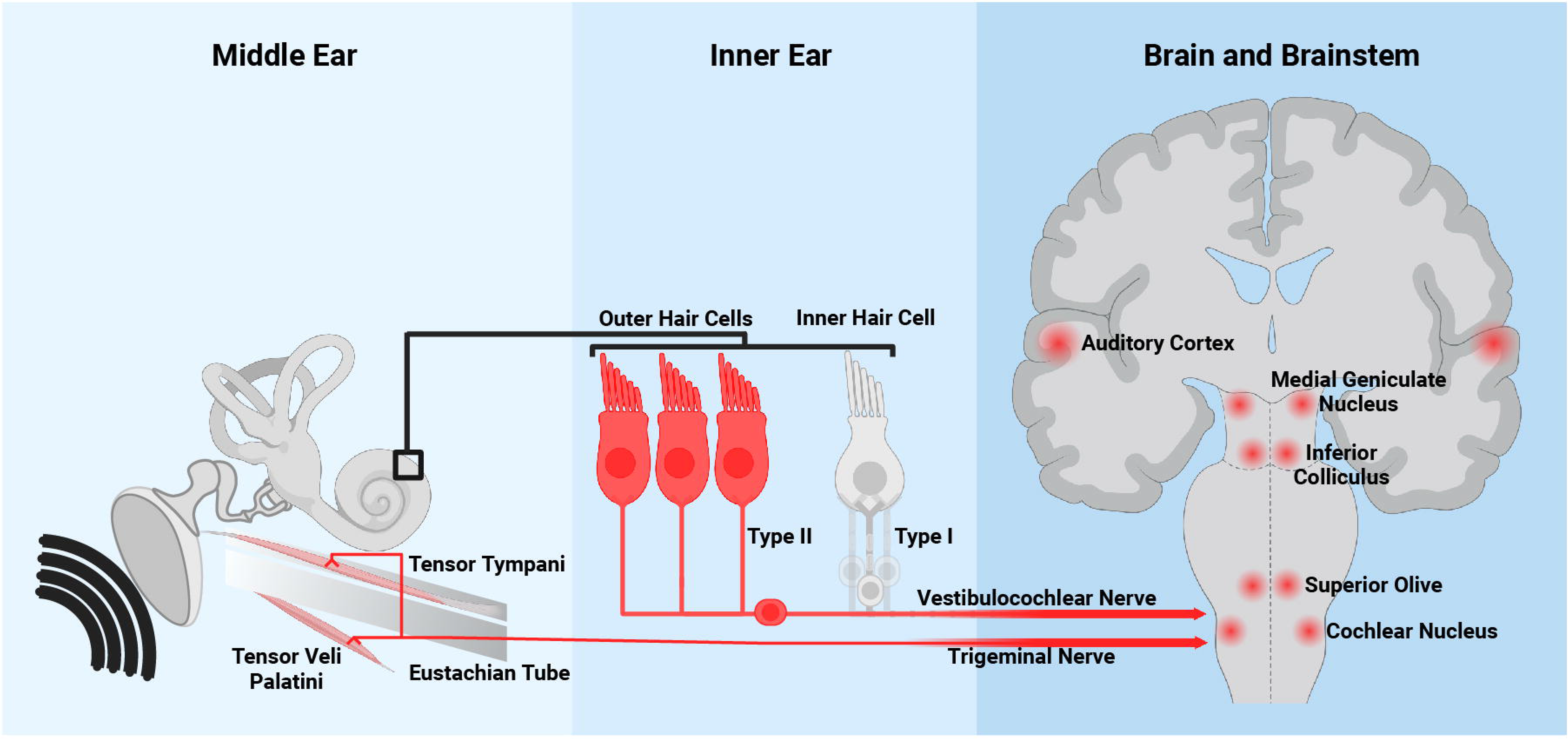
Schematic depicting three potential noxacusis mechanisms. In each model, hypothesized components mediating noxacusis are highlighted in red. (A) In the middle ear model, nociceptive signaling is mediated by the mandibular branch of the trigeminal nerve. Trigeminal nerve afferents innervate the tensor tympani and the tensor veli palatini muscles, both proximal to the middle ear space. (B) In the inner ear model, Type II-afferent nerve fibers synapse with outer hair cells and transduce nociceptive signals to the central auditory system. (C) In the central auditory system, maladaptive hyperactivity in the brainstem and brain potentially mediate hyperacusis.

Existing conceptual frameworks of the biological underpinnings of pain hyperacusis lack empirical evidence from humans, relying largely on theory [12,20] or animal models [1,22]. Consequently, pain hyperacusis remains poorly treated and responds inadequately to existing therapies, highlighting the urgent need for more effective solutions. We surmise that the mechanisms of pain hyperacusis will continue to remain elusive without a thorough understanding of how the condition manifests physically and the types of interventions that can alleviate sound-induced pain. Here, we used a participatory research method to generate a conceptual framework for understanding the most salient symptoms of pain hyperacusis from the perspective of individuals who suffer from the condition. We also identified pharmaceutical and non-pharmaceutical interventions that the participants have tried and the perceived efficacy of each. We aimed to characterize the specific physical and psychosocial consequences of sound-related pain and to generate a preliminary dataset to identify interventions that may provide pain relief. These data will be used to develop targeted clinical trials aimed at identifying effective treatments for pain hyperacusis which, in turn, will aid in uncovering the underlying mechanisms of this condition in humans.

## METHODS

### Participants

This study was approved by the Institutional Review Board of The University of Texas at Dallas (IRB-23-200) and all participants provided written informed consent to participate. A snowball sampling technique was used to recruit prospective participants, and recruitment flyers were posted online on our laboratory website, social media (Facebook, Instagram, and Twitter/X), and a research recruitment registry (ResearchMatch.org). A total of 32 fluent English-speaking adults (age 19-65 years, mean age = 37 years, 11 female) who self-reported that they experience physical pain when they hear sounds participated. All participants had experienced pain hyperacusis for at least one year (range 1 to 27 years). Other self-reported co-morbid audiovestibular, neurological, mental health, and chronic pain conditions are listed in **Table 1**.

**Table 1.** Prevalence of self-reported co-morbid conditions. Thirty-one participants responded to the audiovestibular, neurological, and mental health questions. Chronic pain syndromes and other sensory sensitivities were included in the follow-up survey and thus had 24 total respondents.

| Condition | Prevalence |
| --- | --- |
| ***Audiovestibular*** |  |
| Tinnitus | 96.77% ($n = 30$) |
| Difficulty hearing in noise | 38.71% ($n = 12$) |
| Imbalance or dizziness | 29.03% ($n = 9$) |
| Hearing loss | 25.81% ($n = 8$) |
| Phonophobia | 19.35% ($n = 6$) |
| Misophonia | 16.13% ($n = 5$) |
| Chronic otitis media | 9.68% ($n = 3$) |
| Central auditory processing disorder | 6.45% ($n = 2$) |
| ***Neurological / Mental Health*** |  |
| Anxiety | 48.39% ($n = 15$) |
| Depression | 25.81% ($n = 8$) |
| Head injury | 19.35% ($n = 6$) |
| Post-traumatic stress disorder | 19.35% ($n = 6$) |
| Chronic migraine | 19.35% ($n = 6$) |
| Attention deficit hyperactivity disorder | 9.68% ($n = 3$) |
| Autism spectrum disorder | 6.45% ($n = 2$) |
| William's syndrome | 0.00% ($n = 0$) |
| ***Chronic Pain Syndromes*** |  |
| Temperomandibular joint disorders | 29.17% ($n = 7$) |
| Trigeminal neuralgia | 4.17% ($n = 1$) |
| Fibromyalgia | 4.17% ($n = 1$) |
| Traumatic peripheral nerve injury | 4.17% ($n = 1$) |
| Complex regional pain syndrome | 0.00% ($n = 0$) |
| Diabetic or other metabolic neuropathy | 0.00% ($n = 0$) |
| Autoimmune disorders | 0.00% ($n = 0$) |
| ***Other Sensory Sensitivities*** |  |
| Sight | 41.67% ($n = 10$) |
| No other sensory sensitivities | 37.50% ($n = 9$) |
| Touch | 16.67% ($n = 4$) |
| Smell | 16.67% ($n = 4$) |
| Taste | 4.17% ($n = 1$) |

### Procedures

We first used a modified concept mapping approach to identify the key symptoms and experiences associated with sound-induced pain [3,17]. Concept mapping is a participatory research method that combines qualitative and quantitative techniques to develop a conceptual framework for how stakeholders (here, adults with pain hyperacusis) experience a particular phenomenon. The concept mapping portion of the study consisted of three primary stages (brainstorming, rating, and sorting), outlined in the following sections. We also disseminated a survey to comprehensively characterize the pharmaceutical and non-pharmaceutical interventions that these participants have tried, and the perceived effectiveness of each intervention.

### Concept Mapping Stage 1: Brainstorming (Virtual Focus Group)

The purpose of the concept mapping brainstorming stage is to generate a set of statements that represent the conceptual domain for the topic of interest from the perspective of stakeholders themselves [18]. In the present study, participants (*n* = 26) joined a 60-minute virtual focus group held on the Microsoft Teams platform. During the session, the participants were asked to generate as many statements as possible describing: 1) Characteristics of their sound-related pain, and 2) Symptoms related to sound sensitivity or hyperacusis.

Some participants with severe pain hyperacusis were unable to speak or listen to others’ voices, so we offered multiple accommodations to ensure that all participants had equal opportunity to respond to the focus prompts. Participants were allowed join the focus group with or without audio and we provided instructions for utilizing automatically generated closed captions. Additionally, participants could elect to respond to the focus prompts via either spoken word or text. The Microsoft Teams meeting was recorded so that verbal responses could be transcribed after the session. To facilitate text-based communication, participants received a private link to a Google Docs document to respond to the focus prompts in real time. They were also allowed to enter comments or questions into the Microsoft Teams chat box. All participants had access to the focus prompts during the entire session and were able to freely respond and view other participant responses. Participants were allowed to share additional information about their experiences via email after the focus group.

After the session, the research team reviewed all written and verbal responses. We eliminated redundant ideas and condensed the responses into a set of 92 statements that encompassed all unique ideas generated during the focus group (**Table 2**). The resulting set of 92 statements served as the content for the next two stages of the study (i.e., rating and sorting tasks).

**Table 2.**
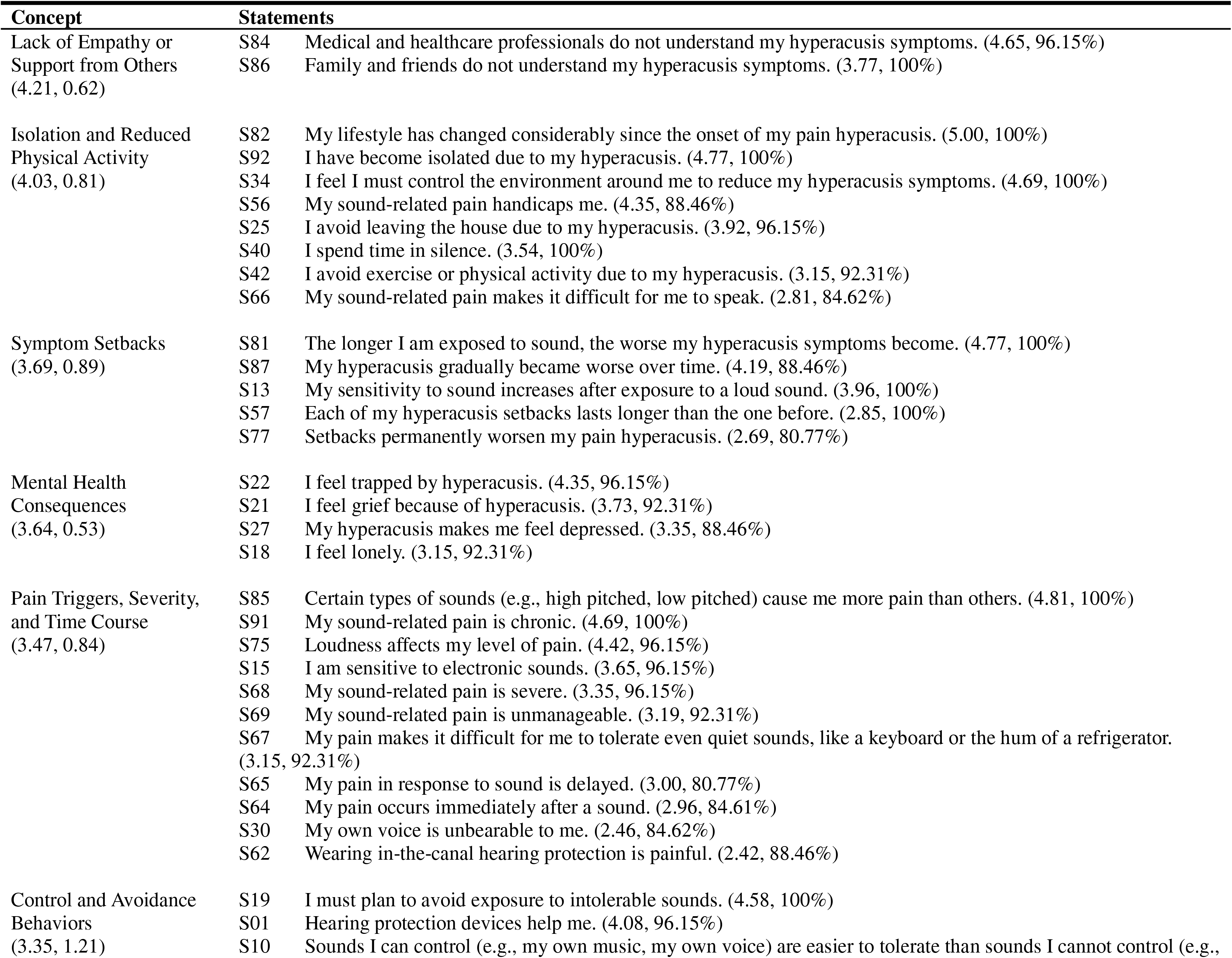
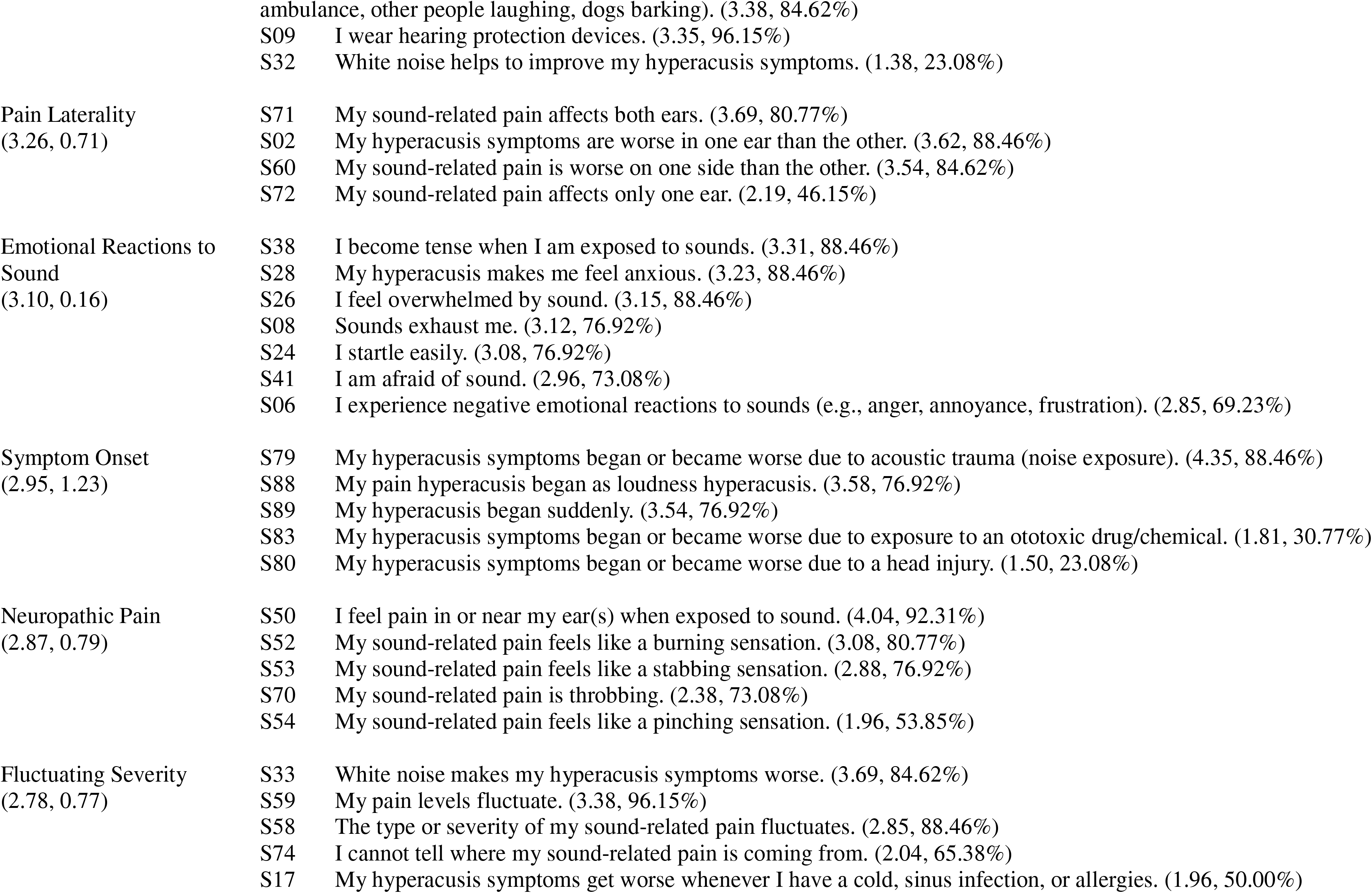
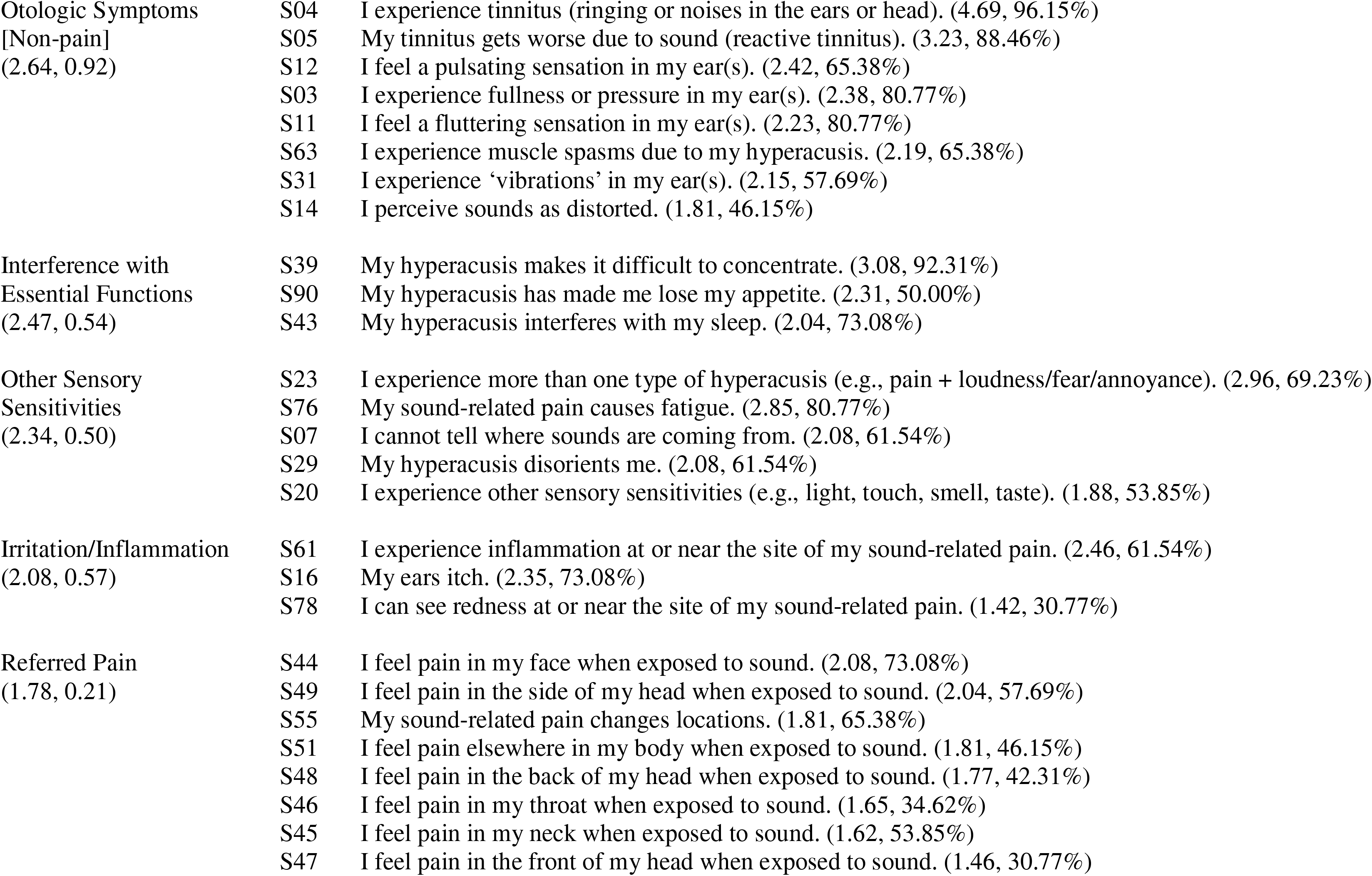
The 17 clusters and the individual statements within each cluster. The average rating and the standard deviation of the ratings for each cluster are in parentheses next to the cluster title. The average rating for each statement and the percentage of participants who endorsed each statement are in parentheses next to each statement. Clusters and statements are ordered from highest to lowest average rating.

### Concept Mapping Stage 2: Rating the Statements

The purpose of the concept mapping rating stage is to allow participants to rate each statement on a dimension of interest to indicate how much the statement embodies their overall experience with the topic [18]. It is possible that some symptoms and consequences of pain hyperacusis are more common than others. Thus, the goal of the rating task in this study was to determine how common it is for an individual with pain hyperacusis to experience each of the symptoms discussed during the focus group. Participants (*n* = 26) completed an online survey where they viewed all 92 statements and rated how well each statement applied to their own personal experience with pain hyperacusis. Twenty of these participants had attended the focus group, and 6 were new participants who were unable to attend the focus group. They received a private link to a Qualtrics survey, where each statement was displayed in random sequence alongside a 5-point Likert scale. Most statements (78 out of 92) were rated on a scale with response choices of *Never, Sometimes, About Half the Time, Most of the Time*, and *Always*. For clarity, a scale with response choices of *Strongly Disagree, Somewhat Disagree, Neither Agree nor Disagree, Somewhat Agree*, and *Strongly Agree* was used for 14 statements [Statement numbers S79-S92; see Table 2] due to their linguistic structure.

### Concept Mapping Stage 3: Sorting the Statements

The purpose of the concept mapping sorting stage is to identify interrelationships, or themes, amongst the statements [18]. Individuals that completed the rating task were invited to complete a follow-up activity where they viewed all 92 statements for a second time and sorted the statements into categories based on perceived similarities between the statements. The goal of this task was to assist the research team in identifying categories and themes within the data. Participants received a private link to view and sort the statements within OptimalSort – Card Sorting software (Optimal Workshop Ltd.). Participants were instructed to organize the statements into categories in a way that made sense to them based on thematic similarity. They were instructed to sort each statement into a category, regardless of whether the statement applied to their own personal situation, and that they should not create a “does not apply” category. They were told that a statement could be placed in its own category if it was unrelated to other statements, but they should not create “miscellaneous” or “other” categories. Once a participant sorted all statements into categories, they were prompted to name each category that they created. Twenty-five participants initiated the sorting task. Of those 25 participants, four did not sort all 92 statements into a category and six created a “does not apply” category despite instructions to the contrary. The categories generated by the remaining 15 participants were used in subsequent analyses. Note that 15 is a sufficient sample size for the sorting stage of a concept mapping study, as the goal of this task is primarily to guide the experimenters in identifying conceptual themes in the data [18].

### Intervention Survey

After the concept mapping portion of the study, we disseminated a survey to determine whether these participants had tried any pharmaceutical or non-pharmaceutical interventions for pain hyperacusis and, if so, whether those treatments were effective at alleviating their sound-induced pain. Twenty-four participants completed the follow-up survey. Participants were asked to indicate whether they have ever used opioid, non-opioid prescription or over-the-counter medications, oral or inhaled cannabinoids (marijuana, THC, and/or CBD), Botox injections, or non-pharmaceutical interventions for pain hyperacusis. If they answered “yes” to any of those questions, they viewed a list of relevant medications or interventions and selected all that they had tried. Each question had an open-ended “Other” option for choices that were not listed. For each intervention selected, participants were asked to indicate whether it helped, hurt, or had no effect on their pain hyperacusis using a 5-point Likert scale with the following options: *Excellent effect (>90% pain relief), Modest effect (75% pain relief), Somewhat effective (50% pain relief), No effect (0% pain relief), Made my noxacusis worse*. For each non-pharmaceutical intervention, participants were asked to describe who administered the treatment (e.g., self-administered, licensed professional).

### Data analysis

Data were analyzed in R (Version 4.2.2) and MATLAB (MathWorks, Inc., Natick, MA). Responses from the sorting task were analyzed using multidimensional scaling and hierarchical cluster analyses. A symmetric 92 × 92 similarity matrix was generated for each participant that indicated which statements they grouped together during the sorting task. The individual similarity matrices were added together to produce a total similarity matrix, which indicated how many participants in the entire sample grouped the same two statements together. The total similarity matrix was transformed into a matrix of Euclidean distances using the MATLAB *pdist* function. Statements that were grouped together more frequently by participants are closer together in Euclidean space (i.e., they have a smaller Euclidean distance) than statements that were grouped together less frequently.

Using the MATLAB *cluster* function, the Euclidean distances were submitted to a hierarchical cluster analysis to group individual statements into clusters of statements that reflected similar concepts [17]. The maximum number of clusters was first selected based on the average number of categories that the participants created (Mean = 12 categories, Range = 3-26 categories). The research team reviewed the statements within the 12 clusters and discussed whether it was appropriate to increase or decrease the number of clusters to better categorize the subjective data [5]. After examining five possible cluster solutions, two of the authors (KNJ and STK) reached a consensus that 17 clusters provided a reasonable level of specificity based on the subjective content matter. The clusters represented 17 different concepts or themes within the data. The authors named each of the 17 clusters with guidance from the category labels that the participants generated during data collection (**Table 2**).

Participants’ rating scores were used to identify the relative importance of each cluster and each statement to the pain hyperacusis experience. This was determined by performing two separate analyses. Point values were assigned to the Likert scale response choices as follows: 1 = *Never*, 2 = *Sometimes*, 3 = *About Half the Time*, 4 = *Most of the Time*, and 5 = *Always* and 1 = *Strongly Disagree*, 2 = *Somewhat Disagree*, 3 = *Neither Agree nor Disagree*, 4 = *Somewhat Agree*, and 5 = *Strongly Agree*. To identify the most salient clusters, the rating scores were averaged across all participants and across all statements within each cluster. Clusters with higher average ratings were interpreted as more common to the lived experience of pain hyperacusis than clusters with lower average ratings.

For individual statements, we also calculated the percentage of participants that provided each Likert scale response. Any response other than “Never” or “Strongly Disagree” was considered an endorsement of the corresponding statement (i.e., the person experiences the symptom at least sometimes or agrees with the statement at least some of the time). Statements that were endorsed by a higher percentage of participants were interpreted as more common to the lived experience of pain hyperacusis than statements that were endorsed by a lower percentage of participants.

## RESULTS

### Concept Mapping

Hierarchical cluster analyses revealed a 17-cluster solution that described the most salient characteristics and consequences of sound-induced pain generated during the focus group (**Table 2**). The 17 clusters arranged in order from highest-to-lowest average rating are: *Lack of Empathy or Support from Others*; *Isolation and Reduced Physical Activity*; *Symptom Setbacks*; *Mental Health Consequences*; *Pain Triggers, Severity, and Time course*; *Control and Avoidance Behaviors*; *Pain Laterality*; *Emotional Reactions to Sound*; *Symptom Onset*; *Neuropathic Pain*; *Fluctuating Severity*; *Otologic Symptoms (Non-pain)*; *Interference with Essential Functions*; *Other Sensory Sensitivities*; *Irritation/Inflammation*; *Referred Pain*; and *Head Sensations*. The average ratings for each cluster ranged from 1.66 for *Head Sensations* (least common across all participants) to 4.21 for *Lack of Empathy or Support from Others* (most common across all participants). Table 2 lists the 17 clusters, the average rating for each cluster, the individual statements within each cluster, the average rating for each statement, and the percentage of participants who endorsed each statement at least sometimes.

Figure 2 shows the percentage of participants who endorsed each statement related to the quality and location of their pain (i.e., clusters describing *Neuropathic Pain* and *Referred Pain*) along with a visual schematic of those data. The pain was most often described as a burning (80.77%), stabbing (76.92%), throbbing (73.08%), or pinching (53.85%) sensation. Although 92.31% of participants reported that the pain can occur in or near their ears, less than half of participants (42.31%) indicated that this was “Always” the case. Many participants reported referred pain in addition to pain in or near the ears, including pain in the face (73.08%), head (side – 57.69%, back – 42.31%, front – 30.77%), throat (34.62%), neck (53.85%), and elsewhere in the body (46.15%; i.e., outside the head and neck region). More than half of participants indicated that their sound-related pain can change locations (65.38%) and that the quality and severity of the sound-related pain can fluctuate (**Table 2**, *Fluctuating Severity* cluster).

**Figure 2.**
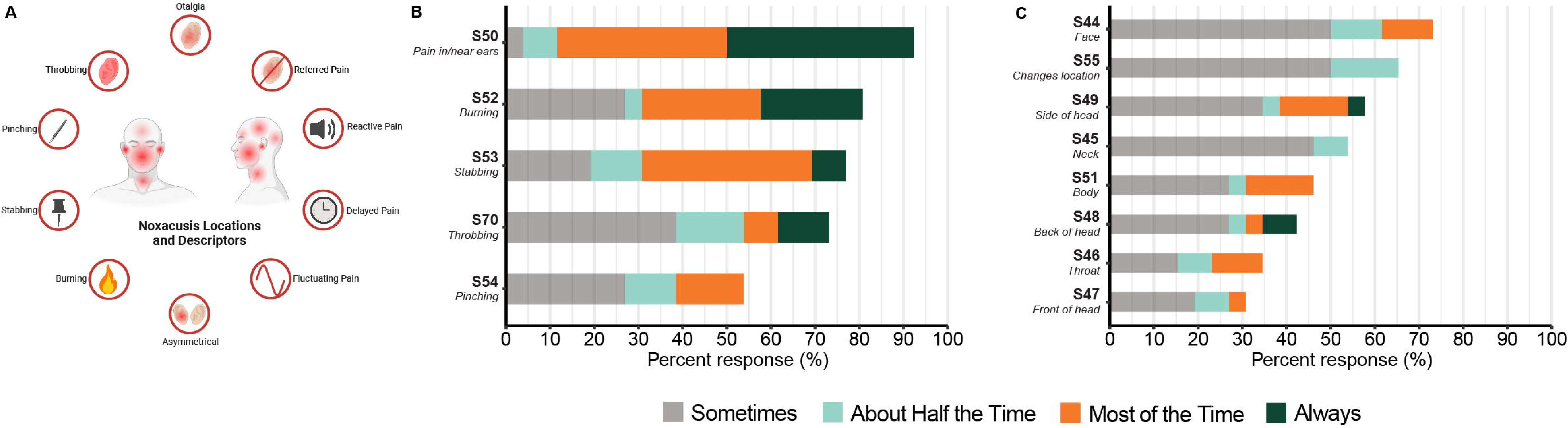
A) Schematic of pain locations and descriptors reported during the focus group and the percentage of participants who endorsed (B) neuropathic pain and (C) referred pain. Statement numbers are displayed on the y-axis along with a key word or phrase representing the content of the statement. Full statements are listed in Table 2.

In addition to the physical characteristics of the pain itself, participants consistently reported experiencing setbacks after sound exposure (**Table 2**, *Symptom Setback* cluster) and described common triggers and factors that influence the severity and temporal characteristics of the pain (**Table 2**, *Pain Triggers, Severity, and Time Course* cluster). We also note that although 80.77% of the participants endorsed bilateral sound-related pain, many reported unilateral pain (46.15%) or symptoms that are asymmetric in severity (84.62%) (**Table 2**, *Pain Laterality* cluster). A smaller, but noteworthy, percentage of participants endorsed symptoms such as inflammation (61.54%), itchiness (73.08%), or redness (30.77%) near the site of their sound-related pain and other head-related sensations such as fullness/pressure (61.54%), migraines (30.77%), dizziness (30.77%), or nausea (26.92%) associated with sound exposure (**Table 2**).

As hyperacusis is often considered to be a disorder of the ear, we were interested in understanding what other otologic and sensory symptoms these individuals may experience (Figure 3; **Table 2**). Almost all participants (96.15%) experience tinnitus (perception of phantom sounds), and their tinnitus is often reactive (88.46%) (i.e., exacerbated by the presence of external sounds) (Figure 3A). Most participants reported additional, non-pain sensations in the ear such as fullness or pressure (80.77%), fluttering (80.77%), pulsing (65.38%), and vibrations (57.69%) and other sensory sensitivities (53.85%) (Figure 3; **Table 1**).

**Figure 3.**
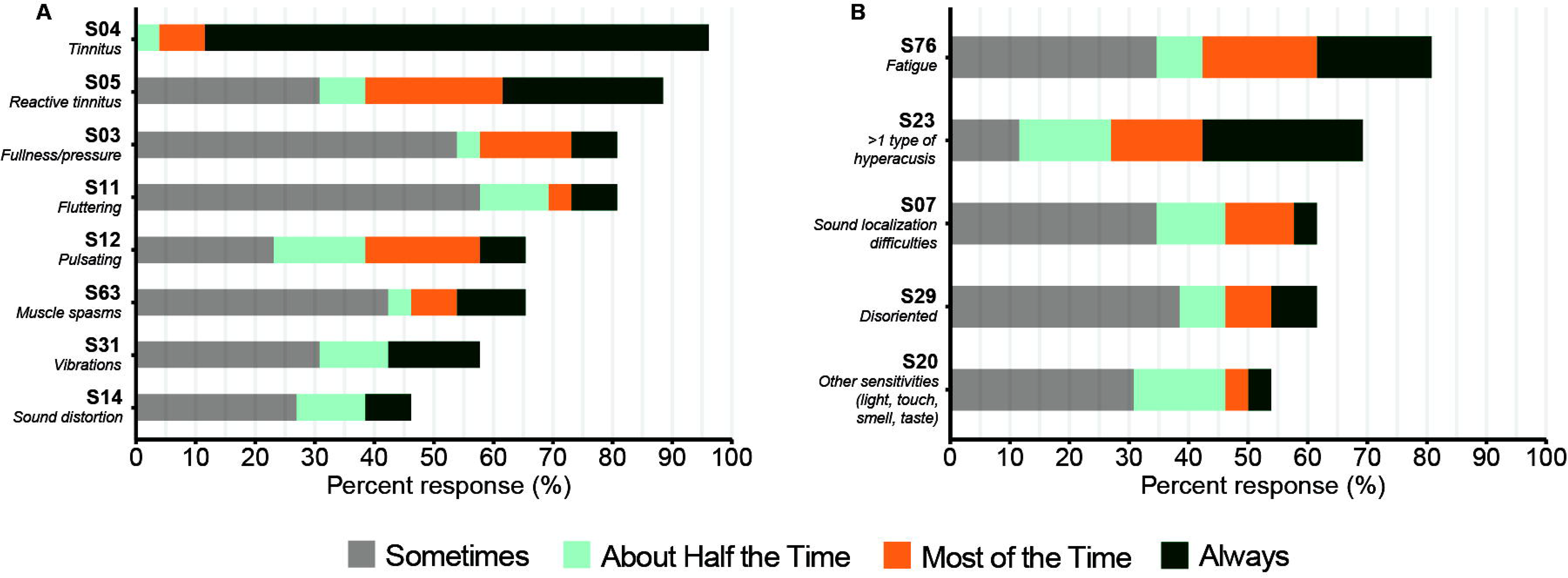
The percentage of participants who endorsed (A) otologic symptoms and (B) sensory sensitivities other than pain. Statement numbers are displayed on the y-axis along with a key word or phrase representing the content of the statement. Full statements are listed in Table 2.

In addition to describing physical and sensory symptoms, participants often discussed the emotional and psychosocial consequences of pain hyperacusis. Clusters related to emotional and psychosocial symptoms were highly rated across participants, suggesting that these experiences may be more consistent than the physical manifestation of the pain itself. Figure 4 shows the percentage of participants who endorsed a lack of empathy or support from others, which was the highest-rated cluster. All participants reported that individuals within their social network do not understand their hyperacusis symptoms, with 100% endorsing this concept for family and friends, and 96.15% indicating that medical and healthcare professionals do not understand their symptoms (Figure 4; **Table 2**).

**Figure 4.**
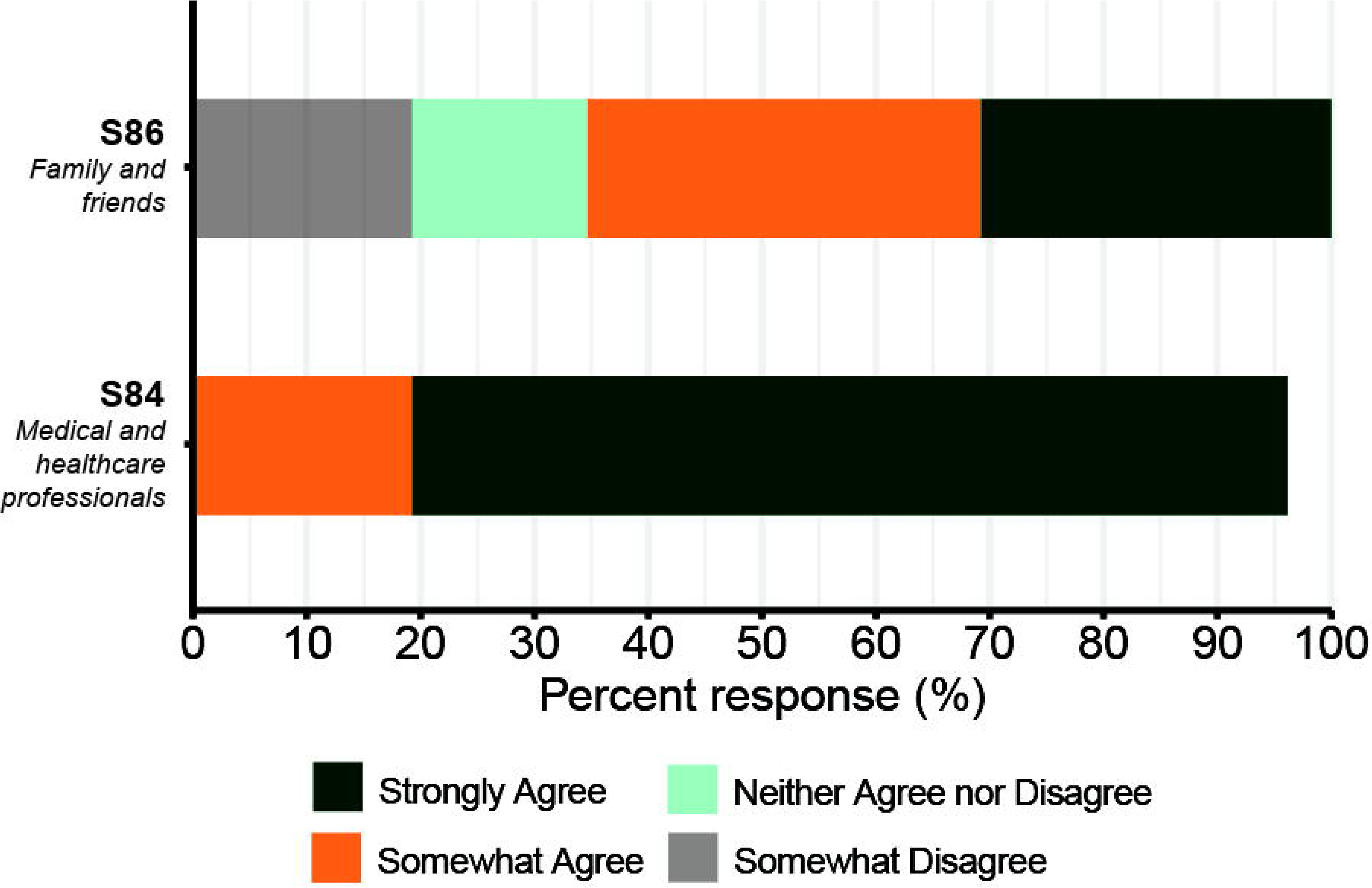
The percentage of participants who endorsed a lack of empathy or support from others. Statement numbers are displayed on the y-axis along with a key word or phrase representing the content of the statement. Full statements are listed in Table 2.

Figure 5 shows the percentage of participants who endorsed each statement within the top two emotional and psychosocial clusters (i.e., *Isolation and Reduced Physical Activity* and *Mental Health Consequences*). Every participant indicated that their lifestyle has changed significantly since the onset of their hyperacusis, with the vast majority endorsing all statements related to isolation and reduced physical activity (range 84.62% to 100%) and mental health concerns (range 88.46% to 96.15%) (Figure 5; **Table 2**). To manage their symptoms, most of the participants use control and avoidance techniques such as wearing hearing protection devices (96.15%) and planning their lifestyle to avoid sound exposure (100%) (**Table 2**).

**Figure 5.**
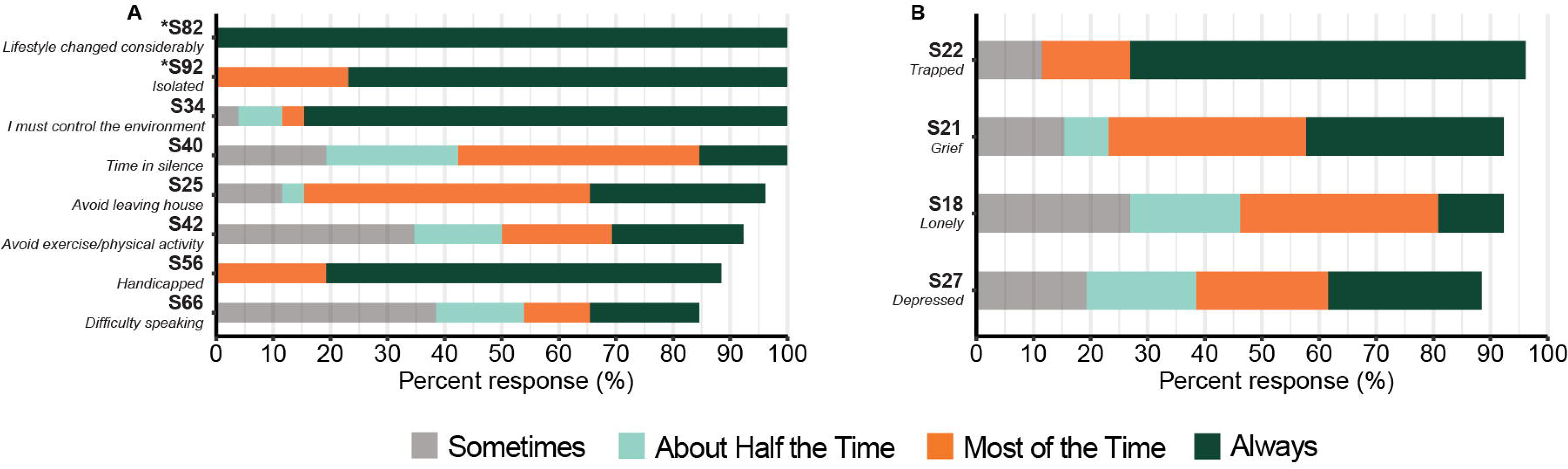
The percentage of participants who endorsed (A) isolation and reduced physical activity and (B) mental health consequences. Statement numbers are displayed on the y-axis along with a key word or phrase representing the content of the statement. Items with an asterisk were rated on a Likert scale ranging from *Strongly Disagree* to *Strongly Agree*. Full statements are listed in Table 2.

### Intervention Survey

Overall, participants unanimously reported pain sensations (Figure 2; e.g., burning, stabbing, pinching) that are consistent with possible damage, disease, or dysfunction of one or more peripheral nerves. We created and disseminated a survey to identify potential treatment response patterns that could shed light on the underlying mechanisms of pain hyperacusis in humans. Participants reported that they have tried a variety of treatments to alleviate sound-induced pain, with most participants (87.50%, *n* = 21) having tried both pharmaceutical and non-pharmaceutical therapies (Figure 6, **Table 3**). The average participant reported using 4.88 different therapies (range 0 to 12), with 58.33% (*n* = 14) trying multiple pharmaceuticals and 66.67% (*n* = 16) trying multiple non-pharmaceutical therapies (Figure 6).

**Figure 6.**
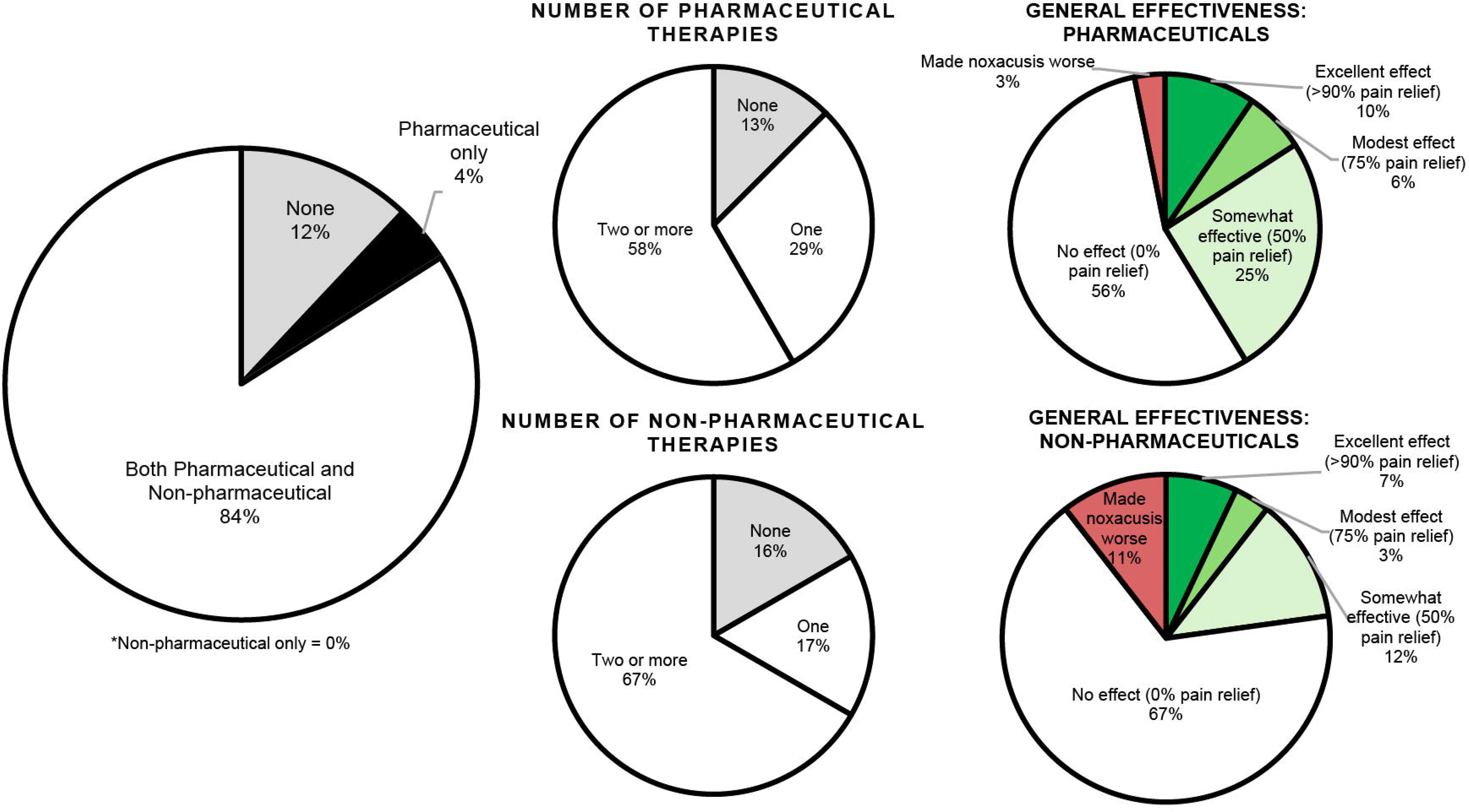
The percentage of participants who tried pharmaceutical or non-pharmaceutical interventions (left panel), the number of therapies tried (middle panel), and the general perceived effectiveness of each type of intervention (right panel).

**Table 3.**
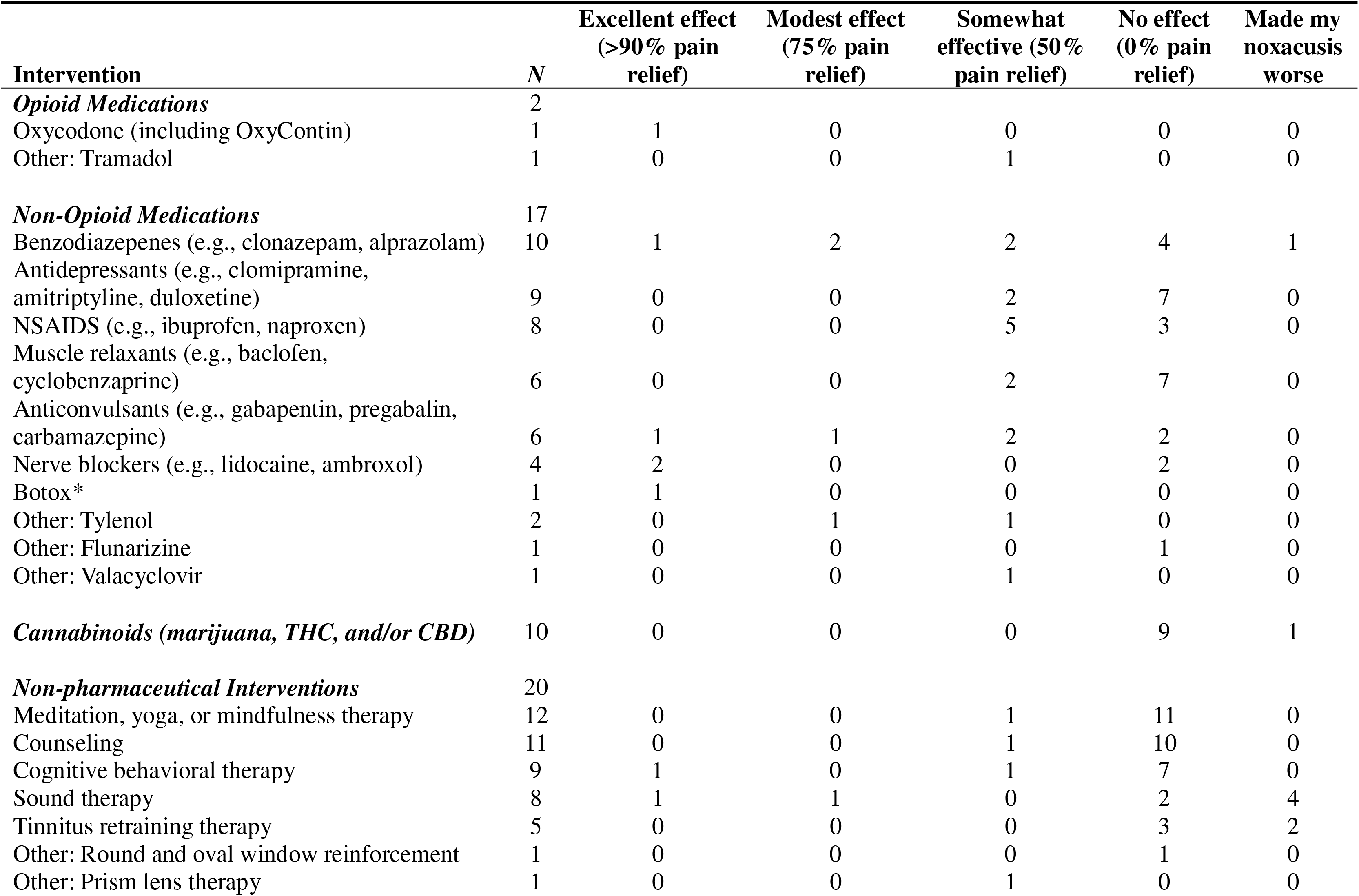
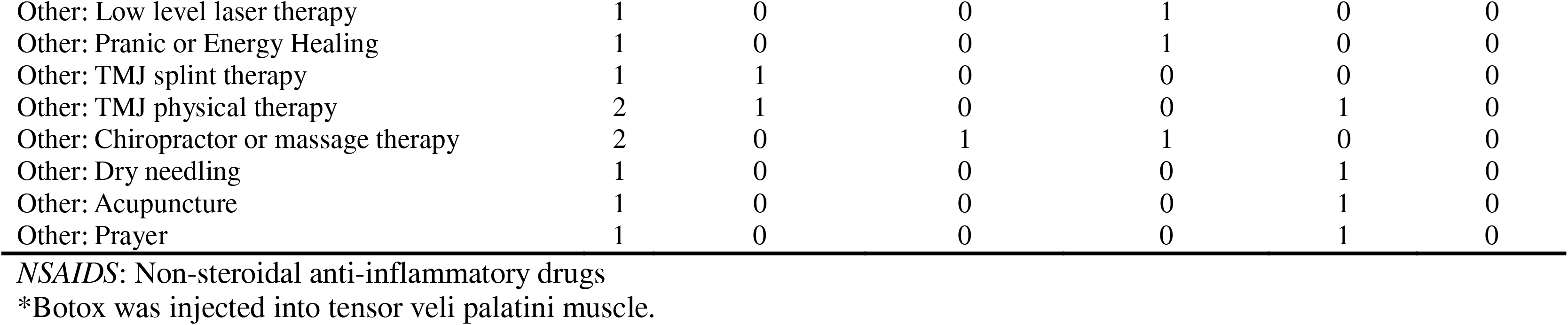
The number of participants (N) who have tried each pharmaceutical and non-pharmaceutical intervention for pain hyperacusis, along with each intervention’s perceived effectiveness.

Those who use pharmaceuticals reported the following drugs, in descending order of frequency: oral or inhaled cannabinoids (e.g., marijuana, THC, and/or CBD) (*n* = 10), benzodiazepines (e.g., clonazepam, alprazolam) (*n* = 10), antidepressants (e.g., clomipramine, amitriptyline, duloxetine) (*n* = 9), non-steroidal anti-inflammatory drugs (NSAIDS; e.g., ibuprofen, naproxen) (*n* = 8), muscle relaxants (e.g., baclofen, cyclobenzaprine) (*n* = 6), anticonvulsants (e.g., gabapentin, pregabalin, carbamazepine) (*n* = 6), nerve blockers (e.g., lidocaine, ambroxol) (*n* = 4), oxycodone (*n* = 1), and Botox injections (*n* = 1). Of the respondents who selected “Other”, two reported using Tylenol, one reported Tramadol (opioid), one reported flunarizine, and one reported valacyclovir.

The perceived effectiveness of these pharmaceutical interventions varied (Figure 6). Out of the 63 effectiveness ratings provided across all pharmaceutical interventions, 55.56% (*n* = 35) of the ratings indicated that the treatment had no effect on pain hyperacusis (0% pain relief) and 3.17% (*n* = 2) indicated that the treatment made their pain hyperacusis worse (3.17%, n = 2). Some patients reported modest-to-excellent effects (75-100% pain relief) from benzodiazepenes (*n* = 3), nerve blockers (*n* = 2), anticonvulsants (*n* = 2), Tylenol (*n* = 1), oxycodone (*n* = 1), and Botox (*n* = 1). The one participant who tried Botox reported that the excellent effect (100% pain relief) was experienced after injection into the tensor veli palatini muscle. That participant previously received Botox injections in the masseter, occipital muscles, and temporal muscles with limited effect. Two other participants reported that they are currently seeking off-label Botox injections for pain hyperacusis. Cannabinoids were consistently rated as ineffective for pain relief (*n* = 9), with one participant indicating that they made their pain hyperacusis worse.

Use and perceived effectiveness of non-pharmaceutical interventions were also highly variable (Figure 6). Respondents who tried non-pharmaceutical interventions reported the following treatments, in descending order of frequency: meditation, yoga, or mindfulness therapy (*n* = 12), counseling (*n* = 11), cognitive behavioral therapy (*n* = 9), sound therapy (*n* = 8), and tinnitus retraining therapy (*n* = 5). The respondents who selected “Other” reported round and oval window reinforcement surgery (*n* = 1), low-level laser therapy (*n* = 1), pranic healing and energy healing (*n* = 1), TMJ-specific splints or physical therapy (*n* = 3), chiropractic or massage therapy (*n* = 2), dry needling and acupuncture (*n* = 1), and prayer (*n* = 1). It was noted that nine participants self-administered at least one of these non-pharmaceutical treatments instead of following a prescriptive protocol administered by a professional.

Non-pharmaceutical interventions were largely ineffective at providing pain relief in our cohort. Out of the 57 efficacy ratings provided across all non-pharmaceutical interventions, most indicated that these treatments had no effect on pain hyperacusis (0% pain relief) (66.67%, *n* = 38) or that these interventions made their noxacusis worse (10.53%, *n* = 6). All six participants who experienced a worsening of their noxacusis indicated that this occurred after therapies that involved a sound exposure component (i.e., sound therapy or tinnitus retraining therapy). Only a few patients reported modest-to-excellent effects (>75% pain relief) of cognitive behavioral therapy (*n* = 1), sound therapy (*n* = 2), TMJ-specific treatments (*n* = 2), and chiropractic or massage therapy (*n* = 1).

## DISCUSSION

### Physical Characteristics of Sound-Induced Pain

We interviewed adults with severe pain hyperacusis about their experiences, and used their responses to generate follow-up surveys that targeted the primary features of sound-induced pain, other pertinent symptoms, and possible interventions. This study differs from previous work [21] in that we focused solely on individuals with severe pain hyperacusis, allowed them to self-describe the symptoms and experiences that are most important to their quality of life, and collected detailed information about interventions and perceived effectiveness. Identifying the type of pain associated with pain hyperacusis is crucial for appropriate pain management, since the therapeutic approaches can vary drastically. Most participants reported physical sensations consistent with neuropathic pain (i.e., burning, stabbing, throbbing, pinching) that can be accompanied by irritation or inflammation of the nerve near the pain site. The pain often fluctuates in severity, location, and quality, and it can occur immediately after a sound exposure, or it can be delayed by several hours. Even though hyperacusis is generally thought to be an ear disorder and many participants had co-morbid otologic symptoms (e.g., tinnitus, aural fullness), they often reported referred pain (i.e. pain that is not restricted to the ear) and non-otologic sensory sensitivities (e.g., light, touch). Many of these symptoms are consistent with a recent report that surveyed individuals with pain hyperacusis and loudness hyperacusis [21].

Most of our participants attributed the onset of their pain hyperacusis to high-intensity noise exposure or ototoxic substances (**Table 2**). Rodent models demonstrate that acoustic overexposure and ototoxicity facilitate a paradoxical increase in sound-evoked and spontaneous neural activity in the central auditory system [1]. This enhanced central gain following sensory loss is believed to underlie the generation of hyperacusis and tinnitus. While basic science work shows abnormally elevated central auditory gain following acoustic injury, there is little-to-no empirical evidence to support central gain as the underlying cause of pain hyperacusis per se. The behaviors evaluated in rodent models may be interpreted as correlates of elevated loudness perception or hyper-reactivity, but it is unclear whether those rodents experience sound-induced pain. It should also be noted that a large body of literature supports a gradual increase in central auditory gain with advancing age [13], yet older adults do not typically develop sound-induced pain as part of the normal aging process. In the present study, most participants reported low efficacy of interventions that are designed to counteract maladaptive gain in the CNS (e.g., sound therapy, tinnitus retraining therapy, benzodiazepines) and seven participants reported that those therapies made their noxacusis worse.

The most pertinent symptoms that our patients report (e.g., neuropathic pain, inflammation, irritation, referred pain, and other otologic symptoms) suggest that the origin of pain hyperacusis may be peripherally mediated as suggested by middle and inner ear models (Figure 1). Approximately 95% of the auditory nerve is comprised of myelinated Type-I neurons that carry acoustic information from the inner hair cells (IHCs) to the cochlear nucleus in the brainstem. The remaining 5% of auditory nerve fibers are unmyelinated Type-II neurons that contact outer hair cells (OHCs). Previous work suggests that Type-II afferents may be involved in auditory nociception, as these fibers have similar morphologic and neurochemical properties to pain-sensing C fibers in the somatic nervous system [4,10,23]. Specifically, both types of fibers can be activated by adenosine triphosphate (ATP), a pain-signaling molecule that is released by damaged tissue, including the OHCs [4,10,23].

Although it is possible that Type-II cochlear afferents could signal tissue damage within the cochlea, damage-evoked activity in these neurons has yet to be linked to perceptual outcomes and it is unclear how this mechanism alone would account for the referred pain and non-pain sensations (e.g., aural fullness, pressure, fluttering, muscle spasms) that our participants report. Instead, it has been hypothesized that overload or damage to the tensor tympani muscle (TTM) in the middle ear can lead to pain that spreads via inflammatory processes to activate the trigeminal nerve and generate symptoms consistent with neuropathic pain and other otologic symptoms [12]. The trigeminal nerve transmits sensory information from the middle ear to the trigeminocervical complex (TCC) in the brainstem and then to the cortex. The TCC integrates a variety of sensory and nociceptive inputs from the middle ear, head, and neck regions, providing a possible explanation for the referred pain and head sensations described by many of our participants. Moreover, loud noises can trigger painful episodes in individuals with trigeminal neuralgia suggesting that the auditory and trigeminal pathways share anatomy [11]. These symptoms are also consistent with the related tonic tensor tympani syndrome, wherein TTM myoclonus triggers pain within and around the ear and other otologic symptoms by irritating the trigeminal nerve, increasing tympanic membrane tension, or altering middle ear ventilation [20]. Chronic, painful irritation of the trigeminal nerve can lead to central pain sensitization, possibly accounting for elevated CNS gain in individuals with pain hyperacusis, if such a phenomenon exists.

Notably, two participants in the present study reported that nerve blockers (e.g., lidocaine, ambroxol) had an excellent effect (>90% pain relief) on their noxacusis, and ambroxol has extensive evidence supporting its use in treating neuropathic pain in conditions such as trigeminal neuralgia, fibromyalgia, and complex regional pain syndrome [16]. Another participant indicated that Botox injections in the tensor veli palatini muscle had an excellent effect (>90% pain relief). The tensor veli palatini is innervated by the trigeminal motor root and may form a functional unit with the TTM to control middle ear pressure [9]. Taken together with existing theories [1,4,10,12,20,23] and other surveys of pain hyperacusis [21], we feel that our results are most consistent with trigeminal nerve involvement. This hypothesis can be clinically tested in the future using locally administered analgesics such as over-the-counter 4% lidocaine ear drops, a common treatment for pain due to acute otitis media in young children [14].

### Emotional and Psychosocial Consequences of Severe Sound-Induced Pain

While the psychosocial consequences of hyperacusis are often acknowledged in the literature [8], few studies have focused on the unique considerations of pain hyperacusis [21], and none have invited participants to self-report the areas of greatest concern and impact. Most striking psychosocial consequences reported in this study were the perceived lack of support and empathy from others, persistent social isolation, and reduced physical activity (**Table 2**). All participants reported that people within their social support network do not understand their condition. Of particular concern was that all but one participant indicated that medical and healthcare professionals do not understand their condition. In fact, the three participants that have not tried any interventions for pain relief wrote that they do not trust healthcare professionals to help manage their pain. This is consistent with a recent study that demonstrated inconsistent clinical practices and insufficient education regarding hyperacusis amongst audiologists [7]. In desperation for support and medical advice, many individuals with pain hyperacusis seek counsel on social media. Avoiding healthcare professionals and seeking counsel from non-medical sources complicates the ability to take a structured approach to treatment.

The present study shows that individuals with pain hyperacusis try many different types of pharmaceutical and non-pharmaceutical interventions that are often self-prescribed. This unstructured treatment approach makes it difficult to identify the treatments and doses that provide pain relief. With the current approach, it becomes challenging to disentangle the myriad factors that may contribute to the efficacy of a specific treatment. Off-label treatment under professional guidance may help to not only relieve pain for individual sufferers, but to generate consistent protocols and clinical datasets that can be leveraged to study and predict treatment success.

### Limitations and Future Directions

There is a critical need for improved education and interdisciplinary approaches to clinical care and research regarding pain hyperacusis. Healthcare professionals should consider appropriate interventions to improve quality of life by managing the numerous physical, social, and emotional burdens associated with pain hyperacusis. Ear specialists need to collaborate with behavioral health and pain management to develop individualized care plans for patients. As is evident from this report, a holistic treatment plan that not only targets the source of the pain itself, but also serves to mitigate mental and physical health consequences in this population is required. Further development of clinically relevant animal models of pain hyperacusis will be critical for advancing our understanding of its pathophysiological mechanisms and for facilitating drug development.

By design, our participants encompassed a small group of individuals with relatively severe pain hyperacusis, and future work will survey a broader and more diverse subset of individuals who experience sound-induced pain. The wide variety of treatments used by our participants precludes systematic analyses of the interrelationships between individual health-related variables, symptom severity, and treatment outcomes. Moreover, we cannot quantify the exact dosages, timing, or duration of treatment administration. Despite these limitations, this work allowed participants to self-generate the most pertinent features of pain hyperacusis that have the greatest impact on their quality of life, and which should serve as primary treatment endpoints in future studies.

## ACKNOWLEDGMENTS

This work was supported by the National Institutes of Health National Institute on Deafness and Other Communication Disorders (NIH NIDCD) K01 DC019647 (PI: Jahn). KNJ reports additional support from the Department of Defense Congressionally Directed Medical Research Program and the American Speech-Language-Hearing Foundation. STK reports support from the American Speech-Language-Hearing Association and the Susan and Jim Jerger Research in Audiology Fellowship. MSY reports support from the NIH National Institute of Diabetes and Digestive and Kidney Diseases (NIH NIDDK) R01 DK134893 (PI: Yousuf). KNJ is a member of the Scientific Advisory Board for Hyperacusis Research Ltd. and serves as the scientific advisor for Hyperacusis Central. The authors declare no other conflicts of interests. We thank Marianne Awad for assisting with data collection, our Hyperacusis Patient Advisory Board for insightful discussions, and the participants who generously offered their time to contribute to this work.

## Data Availability Statement

The conducted research was not preregistered in an independent, institutional registry. Data and analysis code are available from the corresponding author upon reasonable request and completion of an institutional data transfer agreement.

